# Association of early life adversity with internalizing and externalizing behavior across 3 low- and middle-income countries

**DOI:** 10.1101/2025.11.06.25339668

**Authors:** Sheri-Michelle Koopowitz, Richard Shadwell, Nadia Hoffman, Heather J Zar, Giovanni Salum, Pedro M Pan, Gauri Divan, Supriya Bhavnani, Dan J Stein

## Abstract

**Introduction:** Early life adversity (ELA) is associated with increased internalizing and externalizing behavior in high-income cohorts. It is unclear to what extent these associations generalize to low- and middle-income countries (LMICs). We examined the association between ELA and internalizing and externalizing behaviors in children across three LMIC cohorts.

**Methods:** Data from three LMIC cohorts, the Brazilian High-Risk Cohort (BHRC; n = 1111), Sustainable Programme Incorporating Nutrition and Games (SPRING; n = 601), and Drakenstein Child Health Study (DCHS; n = 708) were pooled. Children (7-10 years) were assessed using the Strengths and Difficulties Questionnaire (SDQ). ELA (first 24 months) was assessed using a cumulative index. Ordinary Least Squares linear regression models were run to examine the associations of adversity with internalizing and externalizing behaviors while adjusting for child age, sex, and cohort, and to test interactions with child age and cohort.

**Results:** Measurement invariance did not hold across cohorts, necessitating within-cohort analyses. 2420 children (53.9% boys) were included in analyses. ELA was associated with higher SDQ total scores (B = 1.97, 95% CI: 1.16, 2.78, *p* < .001), internalizing (B = 0.84, 95% CI: 0.41, 1.27, *p* < .001), and externalizing behaviors (B = 1.13, 95% CI: 0.62, 1.64, *p* < .001) in the full sample. Cohort-specific associations were strongest in the BHRC, with attenuated effects in SPRING and DCHS. Girls and older children exhibited fewer behavior problems. Interaction analyses indicated that the associations of adversity with behavior were stronger at younger ages and varied across cohorts.

**Conclusion:** In cohorts from the Global South, while there is an association between early life adversity and psychopathology, there are also important context-dependent differences across cohorts.

## Introduction

Early life adversity (ELA) is a determinant of child neurodevelopment, mental health, and behavior with effects that persist across the lifespan (Berman et al., 2022; Hauser, 2021; Maccari et al., 2014; McLaughlin et al., 2019). ELA encompasses childhood adversities (e.g., abuse, neglect), socioeconomic adversities (e.g., poverty, incomplete education), maternal stress exposures (e.g., traumatic events), and relationship adversities (e.g., family conflict) (Bhopal et al., 2019). Such experiences may disrupt brain maturation during sensitive developmental periods, shaping trajectories of internalizing and externalizing behavior (Berman et al., 2022; Cronholm et al., 2015; Hauser, 2021). Evidence suggests that exposure to multiple, cumulative adversities in the first years of life may have dose-dependent effects (McLaughlin et al., 2021; Oh et al., 2018). This is particularly important in low- and middle-income countries where children experience a variety of adversities in their early years (Bhopal et al., 2019; Walker et al., 2011). Approximately one-third of adult mental disorders are attributable to childhood adversity (McLaughlin et al., 2019), and in children, ELA is associated with internalizing or externalizing behaviors that often precede later psychiatric disorders (Hauser, 2021; Ma et al., 2022).

The majority of research on the association of early life adversity and child behavior has been conducted in high-income countries (Viola et al., 2016), despite most of the world’s children living in low- and middle-income countries (LMICs) (Ma et al., 2022). The extent to which the findings from high-income settings can be generalized elsewhere is unclear. Given the unique and often cumulative risk factors associated with living in low- and middle-income communities (Allen et al., 2016; Lund et al., 2010), it is important to understand how ELA relates to child behavior in low- and middle-income settings.

While several studies have explored the association of early adversity and behavior, most examined isolated adversity exposures or relied on small, cross-sectional samples. By combining data from three independent LMIC cohorts, this study bridged these gaps by utilizing a large, longitudinal dataset that captured multiple adversity domains. Pooled data from three low- and middle-income cohorts (Brazilian High-Risk Cohort, Sustainable Programme Incorporating Nutrition and Games from India, and the Drakenstein Child Health Study from Republic of South Africa) was used to examine whether cumulative early life adversity in the first two years of life was associated with child internalizing and externalizing behaviors. We hypothesized that adversity would predict behavioral difficulties across the cohorts.

## Methods

### Sample and cohorts

Data were collected from children enrolled in three low- and middle-income country cohorts. As each cohort conducted assessments at different time points, outcome data from children aged 7-10 years were utilized in this study.

The Brazilian High-Risk Cohort (BHRC) is a large community school-based study that has followed 2511 children from Brazil since 2010. Psychological, genetic, and neuroimaging data were collected with the aim to investigate typical and atypical trajectories of psychopathology and cognition over development. A two-stage design was used. First, childhood symptoms and family history of psychiatric disorders were assessed in a screening interview, collecting information from 9937 index children at 57 schools in the cities of São Paulo and Porto Alegre, as well as from 45 394 family members. In the second stage, a random subsample (intended to be representative of the community, n = 958) and a high-risk subsample (children at increased risk for mental disorders, based on family risk and childhood symptoms, n = 1554) were selected for further evaluation. These 2511 subjects were evaluated using an extensive protocol, involving one 2-hour home evaluation with the parents, two 1-hour evaluations of the child by a psychologist, and two 1-hour evaluations of the child by a speech pathologist (Salum et al., 2015). Data from n = 1111 BHRC participants were included in this study.

The SPRING (Sustainable Programme Incorporating Nutrition and Games) study is a cluster randomized controlled trial that was conducted in the Rewari district of Haryana, India–the country with the largest number of young children at extreme risk of impaired cognitive and social-emotional development (Lu et al., 2016). 7015 families were enrolled by the surveillance system from 24 clusters, defined as the catchment area of a functional primary health sub-center. Trial outcome measures were assessed in 1443 children at 18 months of age (Kirkwood et al., 2023). SPRING-ELS (Early Life Stress) was a nested sub-study of the SPRING trial evaluating the effects of early adversities and stress on child growth and development (Bhopal et al., 2019). Most recently, through the COINCIDE study, children were followed up at 8 years of age and data from n = 601 participants were included in this study.

The Drakenstein Child Health Study (DCHS) is a multidisciplinary birth cohort study that drew its participants from two peri-urban, relatively stable, low socioeconomic communities in the Western Cape, South Africa (Zar et al., 2015). Pregnant women between 20- and 28-weeks’ gestation were recruited from two primary health care clinics for the main DCHS study. Women were eligible for the study if they were 18 years or older, planned attendance at one of the two recruitment clinics, and intended to remain in the area. During a three-year recruitment period (March 2012 to March 2015), 1225 pregnant women were enrolled into the DCHS antenatally; 88 (7.2%) mothers were lost to follow up antenatally, had a miscarriage or a stillbirth. In total, 1137 women gave birth to 1143 live infants (4 twins and 1 triplet). Data from n = 708 DCHS participants were included in this study.

### Materials

The Strengths and Difficulties Questionnaire (SDQ) is a brief behavioral screening questionnaire that can be used for children between 2 and 17 years old. The SDQ measures 25 attributes designed to measure emotional problems (5 items), conduct problems (5 items), hyperactivity/inattention (5 items), peer problems (5 items), and prosocial behaviors (5 items) using a 3-point Likert scale (Goodman, 1997; Goodman, 2001). Responses for each question range from “not true” being scored 0 points, “somewhat true” being scored 1 point, and “certainly true” being scored 2 points. Total scores can be generated for the individual subscale or an overall total score. To generate a total difficulties score, emotional problems, conduct problems, hyperactivity/inattention, and peer problems are added together (Goodman, 1997). To measure internalizing problems, the emotional problems and peer problems subscales are summed together, while externalizing problems are measured using the conduct problems and hyperactivity/inattention symptoms subscales (Goodman et al., 2010). The SDQ has acceptable reliability and validity across Western and non-Western settings (Muris et al., 2003).

To facilitate comparisons across cohorts, adversity scores were standardized within each sample. Using the SPRING cohort adversity variable (Bhopal et al., 2019) as a template, the individual cohorts created an adversity variable using equivalent measures that reflected the nuances of the individual cohorts. The SPRING adversity variable was chosen due to the focus on early life adversity – adversity taking place within the first year of the child’s life (Bhopal et al., 2019). This variable measured 22 adversities across 4 groups of adversities: childhood adversities, socioeconomic adversities, maternal stress adversities, and relationship adversities (Bhopal et al., 2019). Adversity scores ranged from 0–22 in the SPRING cohort and from 0–12 in the BHRC and DCHS cohorts. Table 1 highlights the adversity variables per cohort, and Figure 1 demonstrates the adversity distribution per cohort. Where exact item matches were not possible, cohort-specific variables were harmonized to reflect the same adversity domains; any exposure was then dichotomized (0 = no ELA exposure, 1 = any ELA exposure) for the primary analyses.

**Figure 1.**
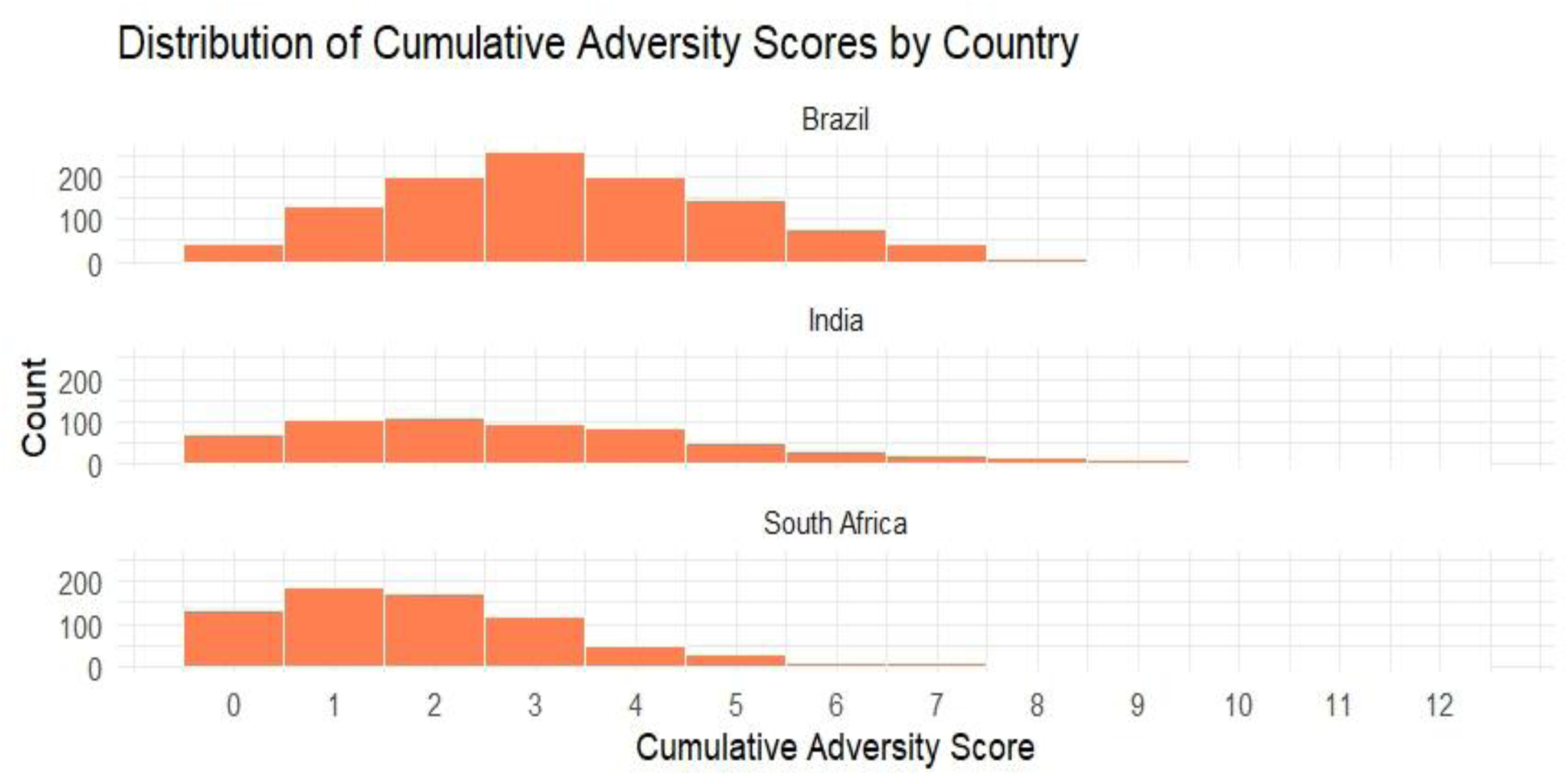
Distribution of Cumulative Adversity Scores by Country.

**Table 1.**
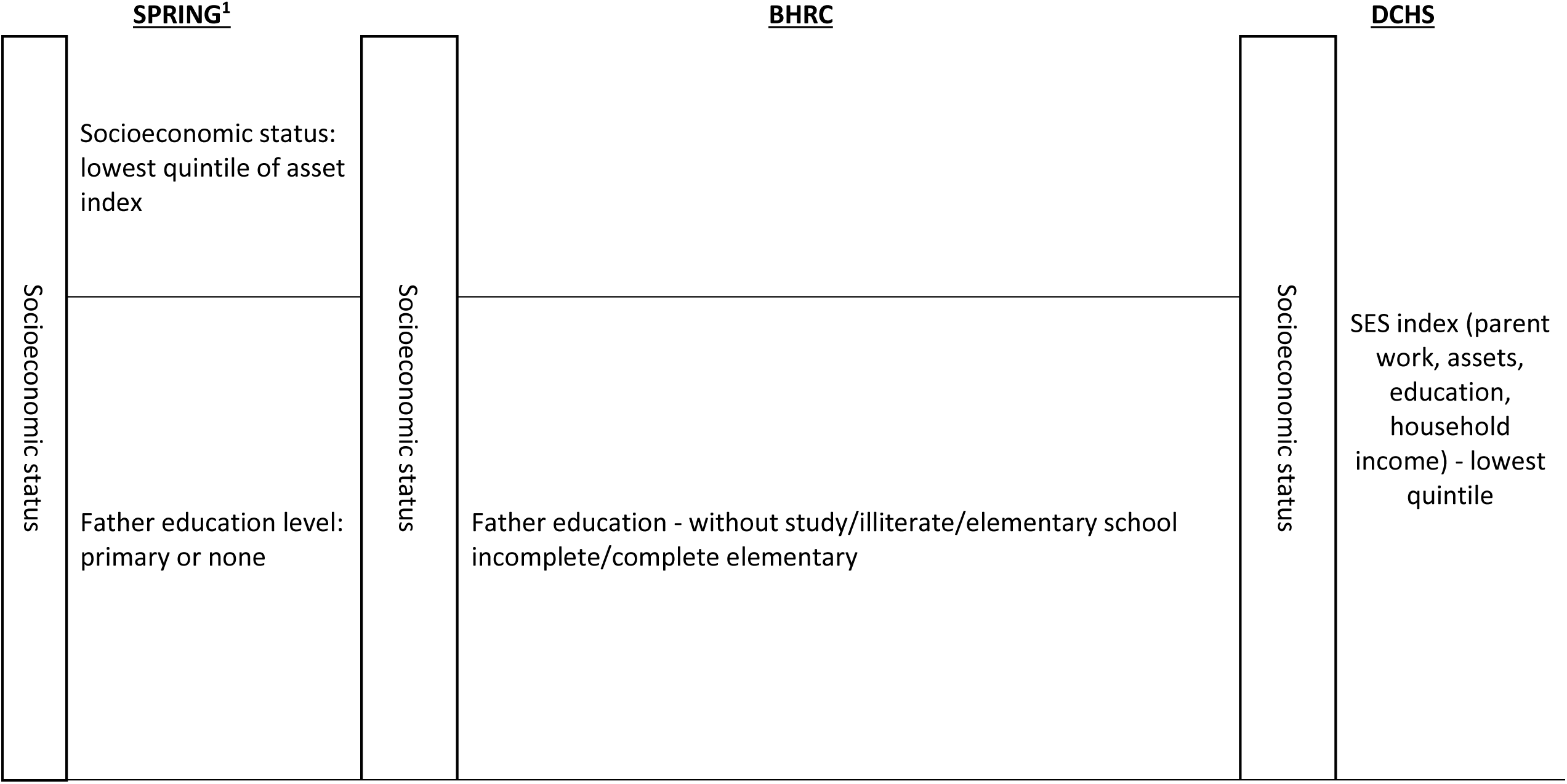
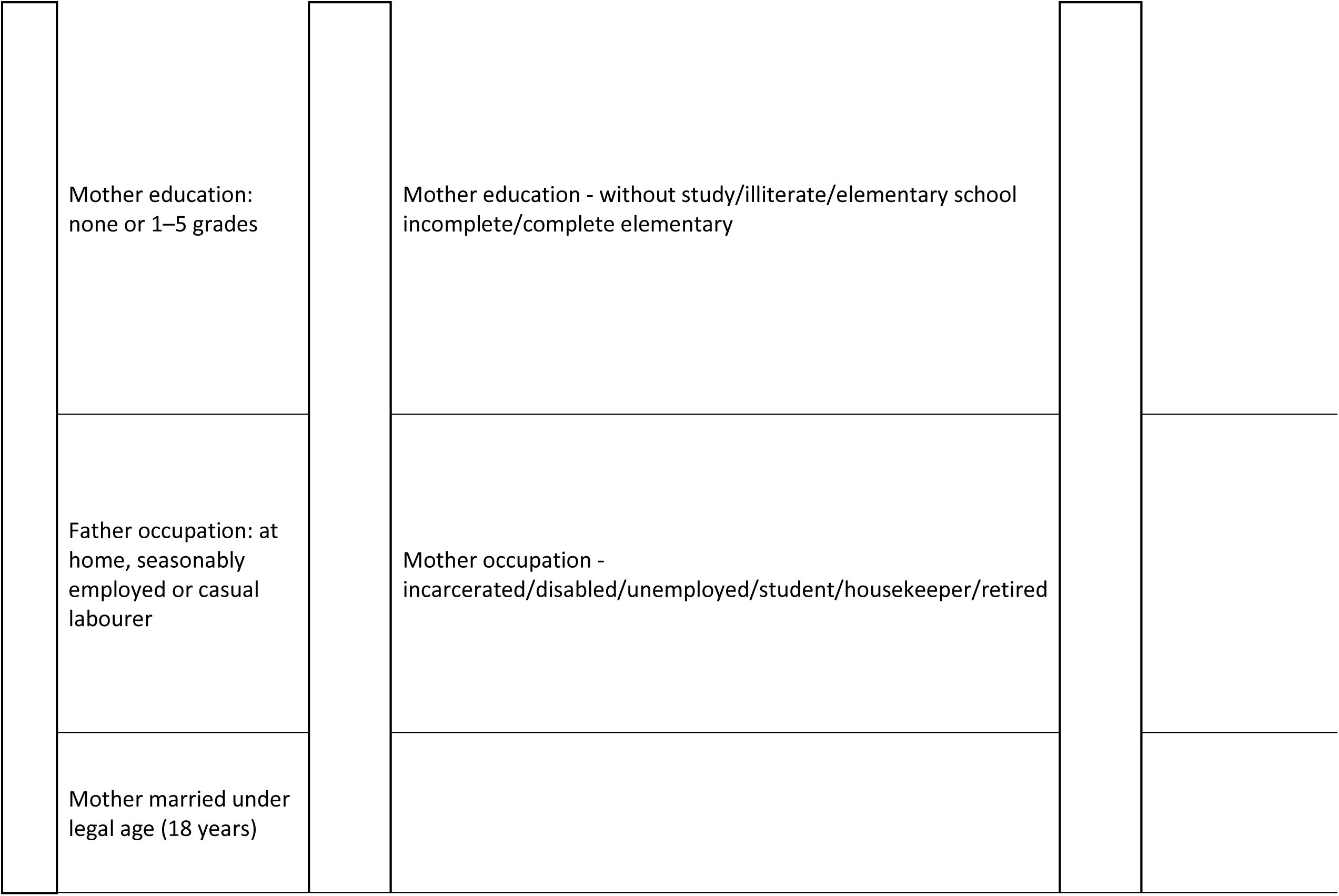
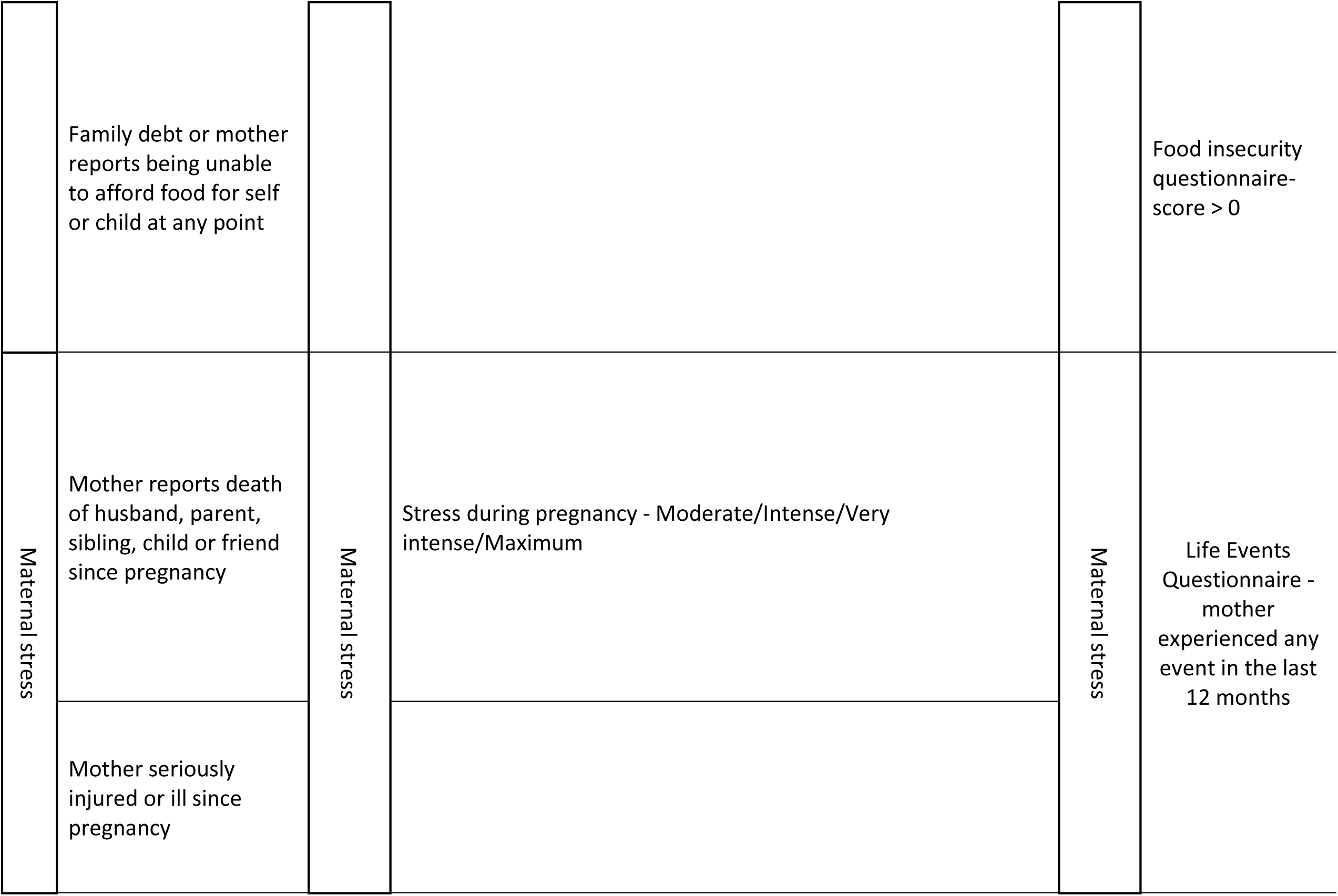
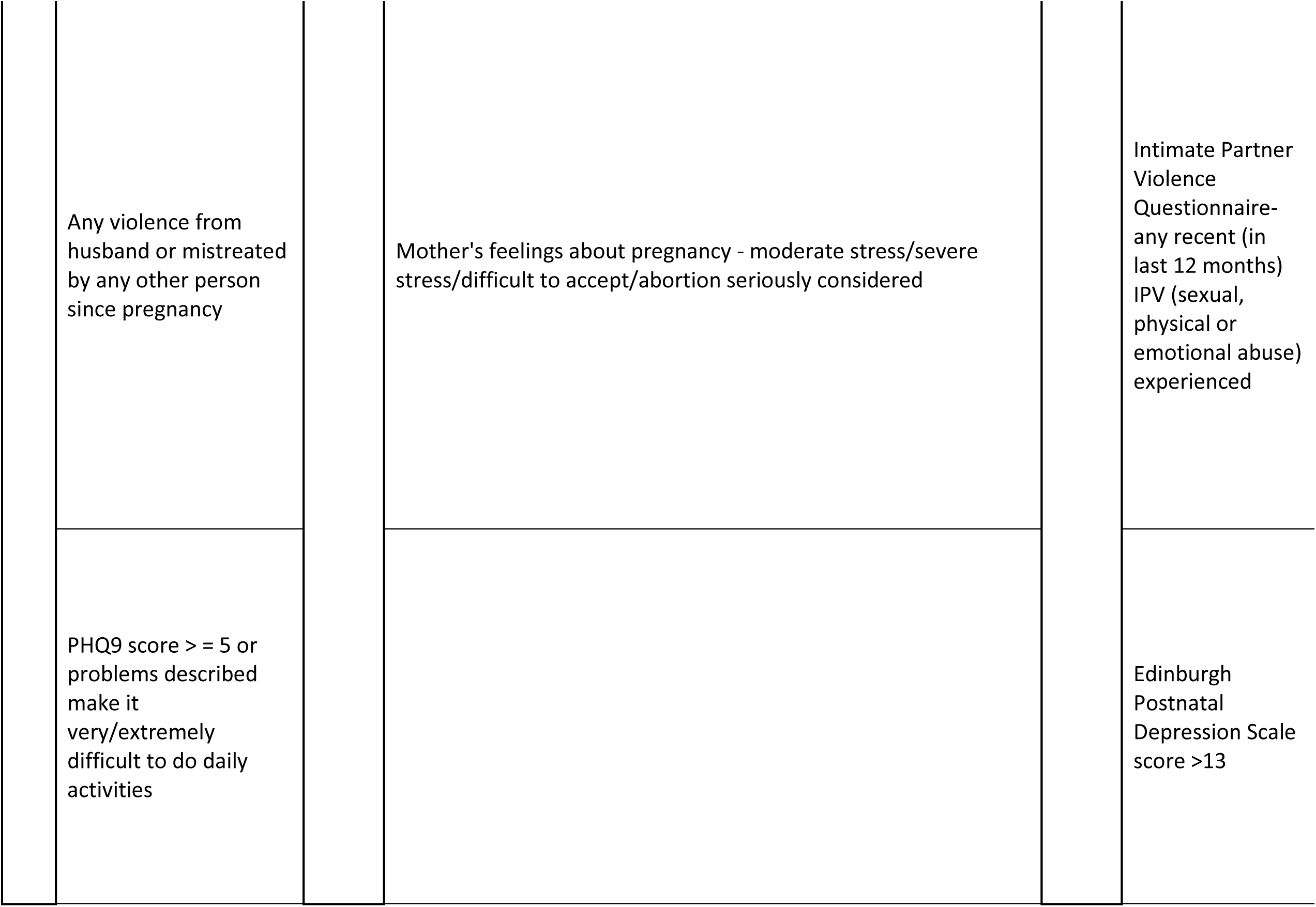
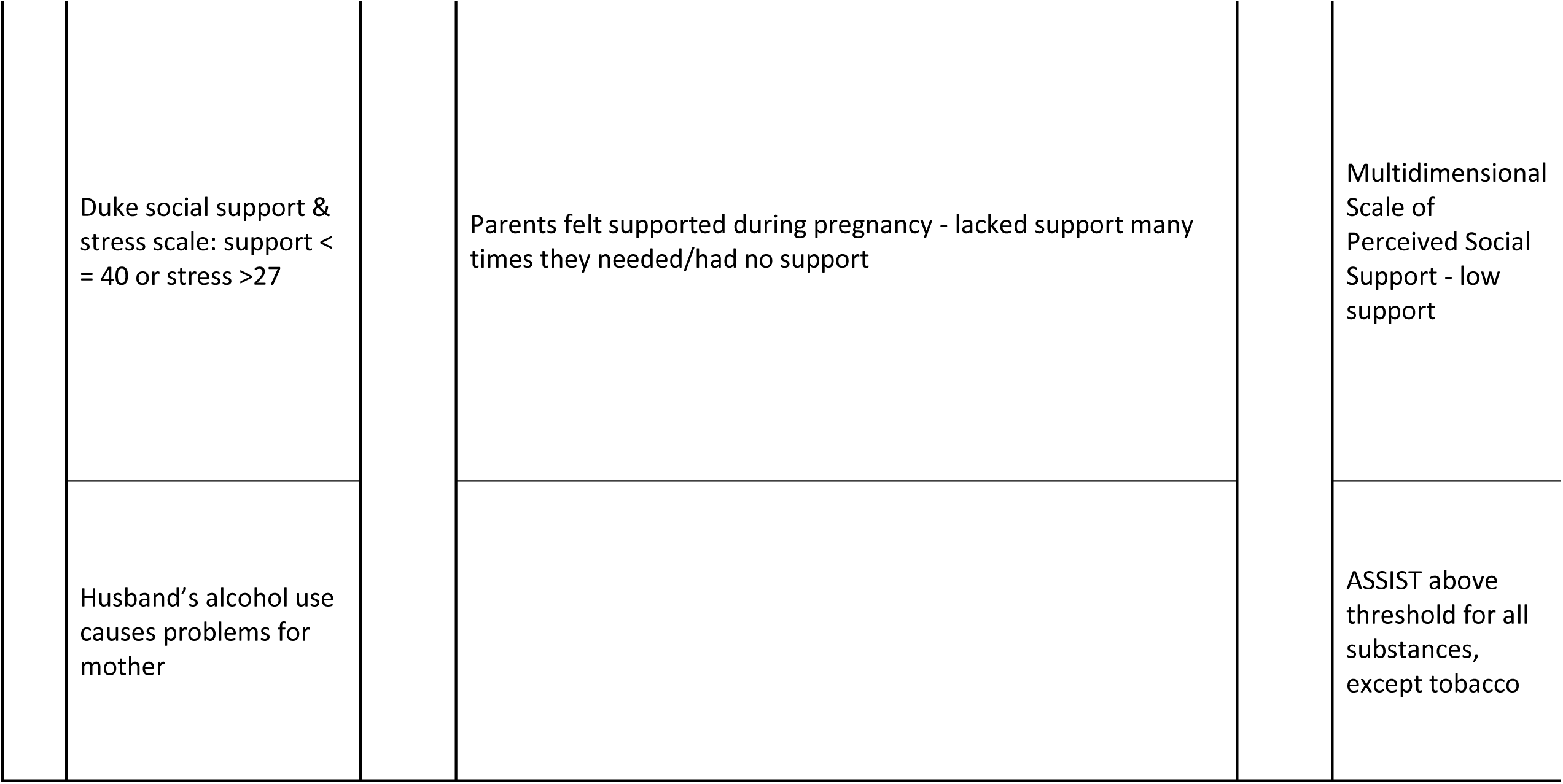
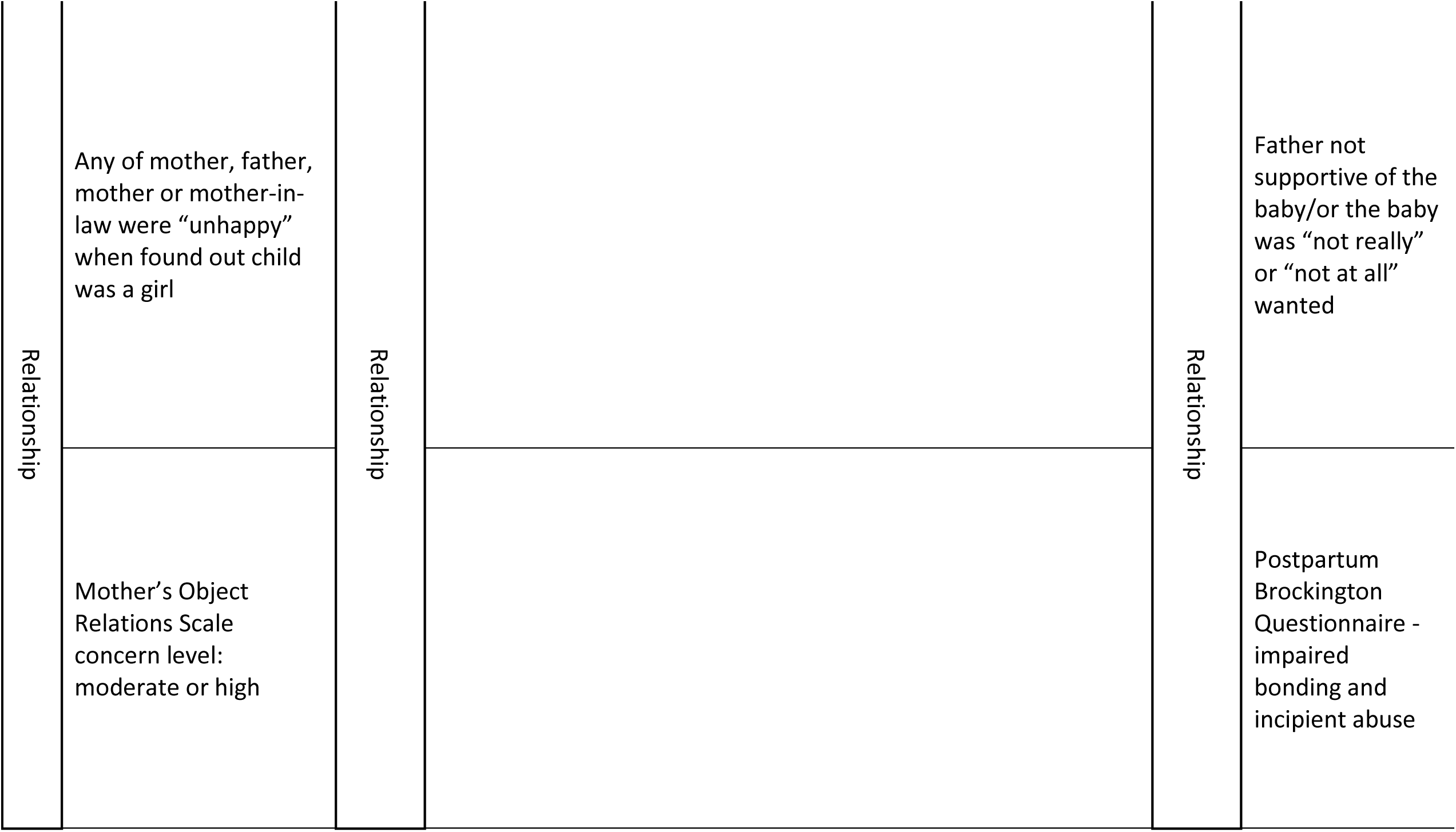
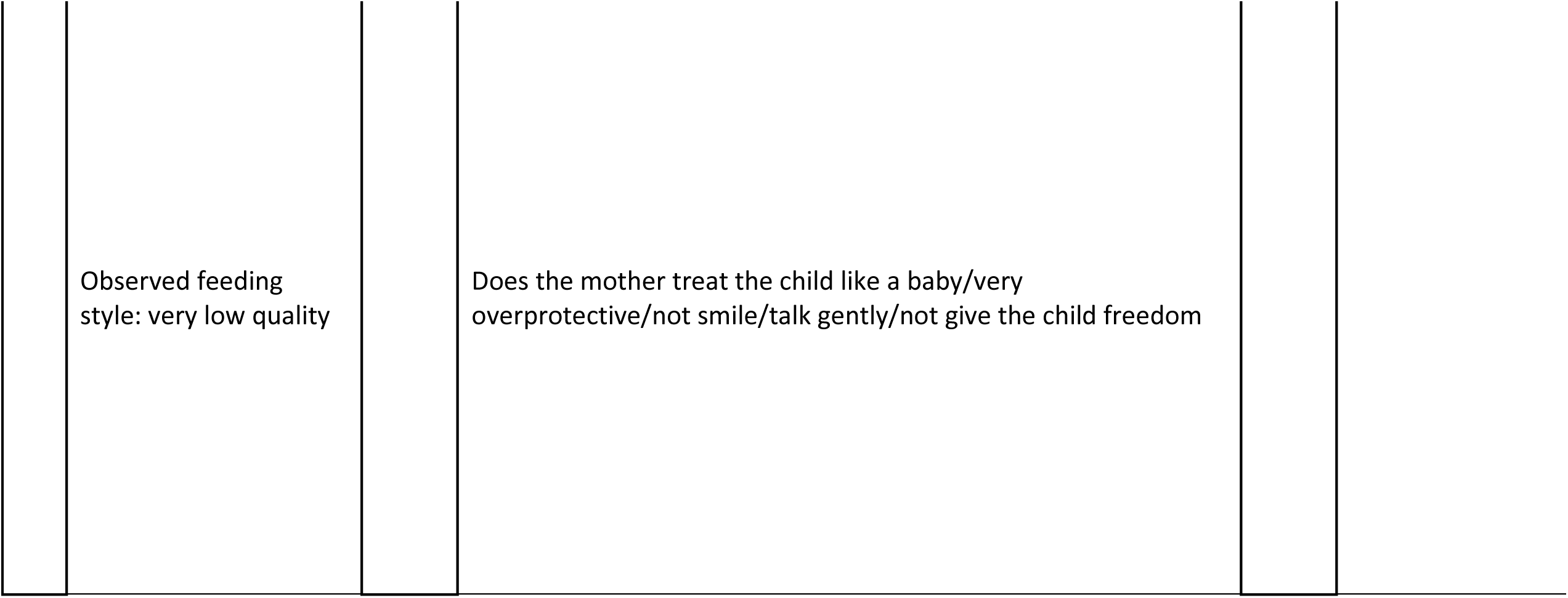
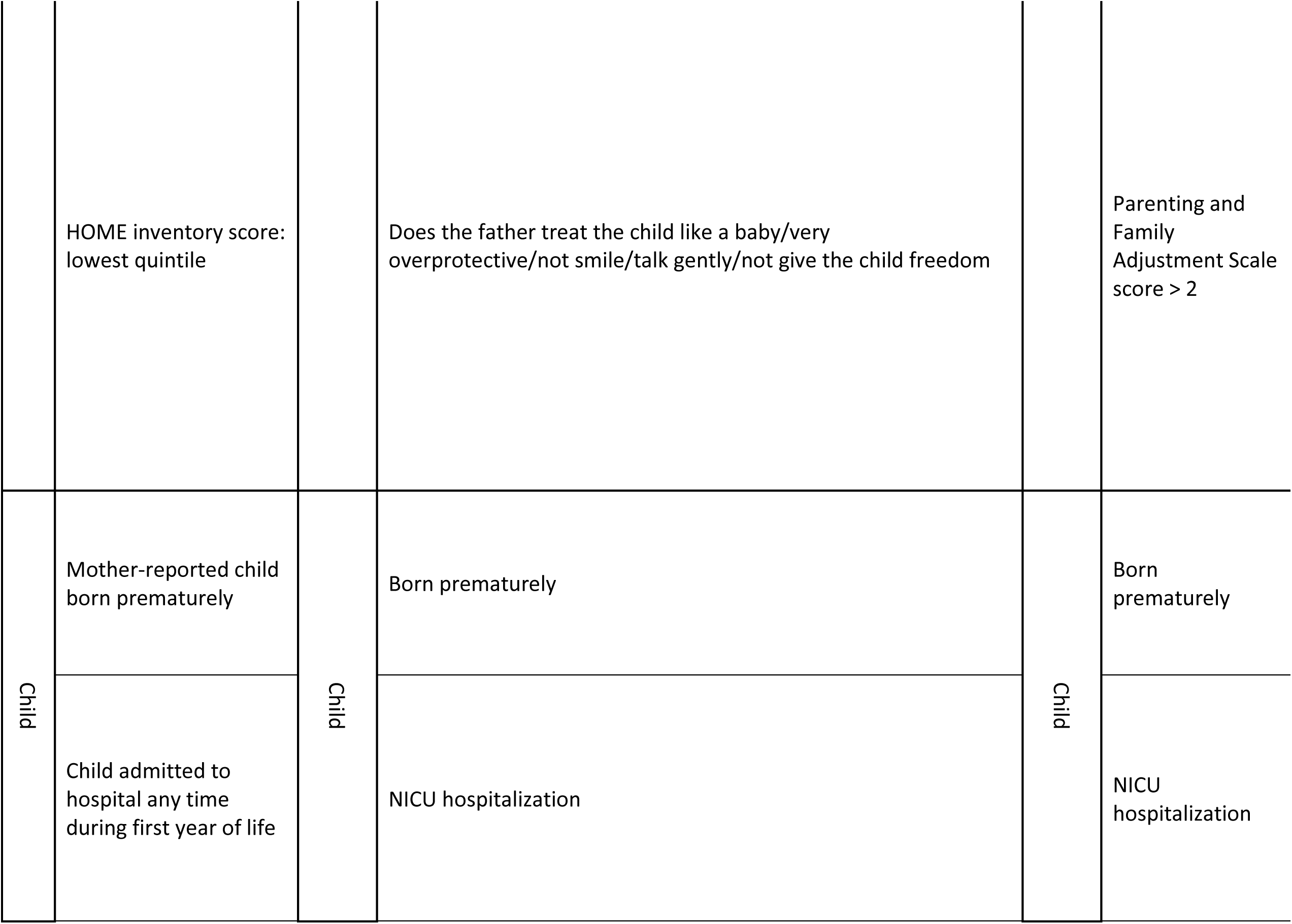
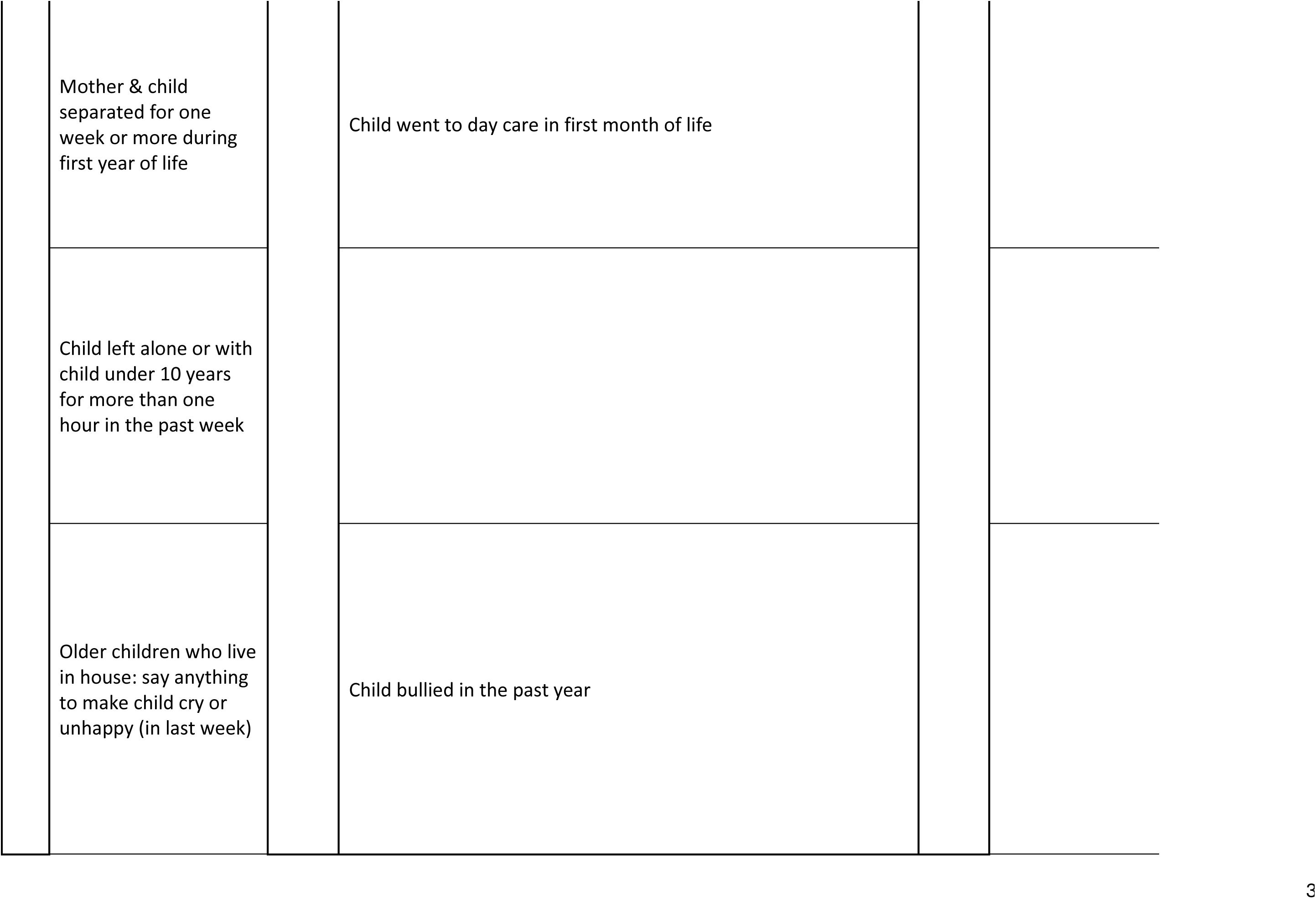
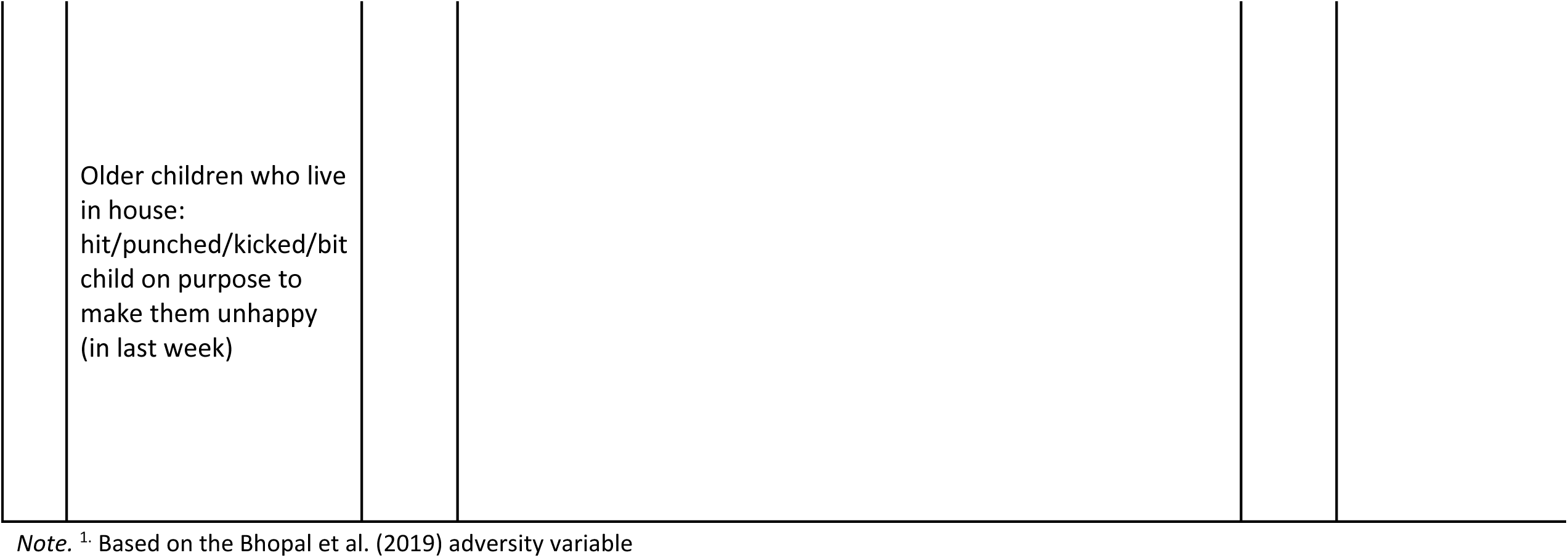
Cohort adversity variables.

### Data analysis

Data were analyzed using *R* (R Core Team, 2021). The association between early life adversity and internalizing and externalizing behaviors was examined using multiple linear regression models, controlling for child age, child sex, and cohort. SDQ measurement invariance (MI) was used to test where the optimal measurement model was equivalent across the cohorts. Measurement invariance indicates the extent to which a measure is interpreted in the same way across different groups of individuals, and without measurement invariance, the measure may be subject to measurement errors (Holden et al., 2020). To test measurement invariance, R’s *lavaan* was used. If measurement invariance did not hold, we planned to conduct within-cohort analyses using: 1) multiple linear regression models with child age and child sex as covariates, and 2) linear models with interaction terms included. We report unstandardized coefficients (B) in-text and standardized coefficients (β) in the tables.

### Ethical considerations

The BHRC obtained ethical approval from the Brazilian National Ethics in Research Commission (CAAE 13852413010015327 and 74563817.7.1001.5327). For the Screening, Wave 0, Wave 1, and Wave 2 stages written consent was obtained from the parents of the participants, as well as from those participants who were able to read, write, and understand the written consent. For those who could not, verbal agreement was obtained. The SPRING study obtained ethical approval from the Sangath Institutional Review Board (GD_2019_55, 28 August 2019 and GD_2022_77, 4 August 2022). Written informed consent was taken from parents or guardians of all participating children, and verbal assent from children above 7 years age. The DCHS was approved by the Faculty of Health Sciences Human Research Ethics Committee, University of Cape Town (401/2009) and by the Western Cape Provincial Health Research committee. Mothers provided informed consent at enrolment and were re-consented annually, and consent was obtained in the mother’s preferred language: English, Afrikaans or isiXhosa. The current study was approved by the Faculty of Health Sciences Human Research Ethics Committee, University of Cape Town (640/2024). Each cohort obtained individual ethical approval from the relevant institutions.

## Results

### Measurement Invariance

SDQ measurement invariance was tested sequentially across cohorts. The configural model (no equality constraints) provided a borderline baseline fit (TLI = 0.888). Imposing metric invariance constraints significantly worsened the model fit compared to the configural model, χ²(32) = 729.8, *p* < .001, with TLI decreasing to 0.884. Further constraining the model to scalar invariance also resulted in a significant decrease in fit relative to the metric model, χ²(32) = 810.1, *p* < .001, with TLI decreasing to 0.854. As scalar invariance was not achieved, pooled effect estimates may be biased; therefore, we present within-cohort regression results in addition to pooled analyses.

### Sample characteristics

Sample characteristics can be found in Table 2. A total of *N* = 2420 participants were included in the analyses with 53.9% boys. The mean child’s age was 8.3 years (*SD* = 0.7).

**Table 2.** Sample characteristics.

|  | Full sample | BHRC | SPRING | DCHS |  |  |
| --- | --- | --- | --- | --- | --- | --- |
|  | N = 2420 | n = 1111 | n = 601 | n = 708 | *F or $\chi^2$* | *p* |
| Age | 8.3 (0.7) | 8.6 (0.8) | 7.9 (0.3) | 8.03 (0.2) | 352.5 | < .001 |
| Male % | 53.8% | 56.3% | 54.6% | 49.7% | 7.8 | .02 |
| SDQ total score | 10.3 (7.5) | 14.8 (7.8) | 4.1 (3.5) | 8.5 (4.5) | 655.7 | < .001 |
| Internalizing | 4.4 (3.9) | 6.8 (4.1) | 1.8 (1.9) | 2.9 (2.3) | 608.1 | < .001 |
| Externalizing | 5.9 (4.5) | 7.9 (4.9) | 2.3 (2.2) | 5.6 (2.9) | 437.1 | < .001 |

*Note.* SDQ denotes Strengths and Difficulties Questionnaire. F tests for continuous variables and Chi Square for categorical variables

The Brazilian High-Risk Cohort included a total of n = 1111 participants in this study. 1400 children (aged 11-14 years) did not meet the 7–10-year age range criteria and were excluded from analysis. The mean child age was 8.6 years (*SD* = 0.8), and over half of this cohort (56.3%) were boys.

The Sustainable Programme Incorporating Nutrition and Games cohort included a total of n = 601 participants in this study. No participants were excluded due to missing data. The mean child age was 7.9 years (*SD* = 0.3), and over half of this cohort (54.6%) were boys.

The Drakenstein Child Health Study cohort included a total of n = 1143 participants in this study. After excluding n = 435 participants due to incomplete or missing data, analyses included n = 708 participants. The mean child age was 8.0 years (*SD* = 0.2), and under half of this cohort (49.7%) were boys.

The cohorts differed significantly in child age (*F*(2, 2417) = 352.5, *p* < .001), with BHRC a significantly older cohort than both SPRING (*p* < .001) and DCHS (*p* < .001). The cohorts differed in child sex (*χ^2^* = 7.8, *p* = .02), with more boys in BHRC than SPRING and DCHS.

### Primary findings

Table 3 highlights the standardized estimates (β) for the models exploring early life adversity and behavior. Exposure to early life adversity was significantly associated with higher total SDQ scores (B = 1.97, 95% CI: 1.16, 2.78, *p* < .001). Girls had lower total SDQ scores than boys (B = –0.95, 95% CI: –1.43, –0.47, *p* < .001). Children from India (B = – 10.79, 95% CI: –11.45, –10.13, *p* < .001) and South Africa (B = –6.14, 95% CI: –6.77, –5.51, *p* < .001) had lower scores than children from Brazil. The final model explained 36% of the variance in SDQ scores (Adjusted *R²* = .361, *F*(5, 2414) = 274.1, *p* < .001).

**Table 3.** Standardized regression coefficients for associations between adversity and behavior outcomes.

|  | SDQ total score |  | Internalizing |  | Externalizing |  |
| --- | --- | --- | --- | --- | --- | --- |
| | $\beta$ [95% CI] | p | $\beta$ [95% CI] | p | $\beta$ [95% CI] | p |
|  |  | value |  | value |  | value |
|  |  | < |  | < |  | < |
| Adversity | 0.08 [0.05, 0.11] | .001 | 0.07 [0.03, 0.10] | .001 | 0.08 [0.04, 0.11] | .001 |
|  |  |  |  |  | -0.05 [-0.09, - |  |
| Age (years) | -0.03 [-0.06, 0.01] | .129 | -0.00 [-0.04, 0.04] | .979 | 0.01] | **.017** |
|  | -0.13 [-0.19, - | < |  |  | -0.19 [-0.26, - | < |
| Sex (Girl) | 0.06] | **.001** | -0.03 [-0.09, 0.04] | .437 | 0.12] | **.001** |
|  | -1.43 [-1.52, - | < | -1.28 [-1.37, - |  | -1.31 [-1.40, - | < |
| Country (India) | 1.35] | **.001** | 1.19] | **<.001** | 1.21] | **.001** |
| Country (South | -0.82 [-0.90, - | < | -0.98 [-1.07, - |  | -0.52 [-0.61, - | < |
| Africa) | 0.73] | **.001** | 0.90] | **<.001** | 0.43] | **.001** |

Note. Coefficients are standardized regression estimates ( $\beta$ ).

Exposure to early life adversity was significantly associated with higher internalizing scores (B = 0.84, 95% CI: 0.41, 1.27, *p* < .001). Children from India (B = –4.97, 95% CI: – 5.32, –4.62, *p* < .001) and South Africa (B = –3.83, 95% CI: –4.16, –3.5, *p* < .001) had significantly lower internalizing scores than children from Brazil. The adjusted model explained 34% of the variance in internalizing scores (Adjusted *R²* = .338, *F*(5, 2414) = 247.6, *p* < .001).

Exposure to early life adversity was associated with higher externalizing scores (B = 1.13, 95% CI: 0.62, 1.64, *p* < .001). Each additional year of age was associated with lower externalizing scores (B = –0.31, 95% CI: –0.56, –0.06, *p* = .017), and girls had significantly lower scores than boys (B = –0.85, 95% CI: –1.15, –0.55, *p* < .001). Compared with children from Brazil, children from India (B = –5.82, 95% CI: –6.23, –5.40, *p* < .001) and South Africa (B = –2.31, 95% CI: –2.7, –1.19, *p* < .001) had significantly lower externalizing scores. The adjusted model explained 28% of the variance in externalizing scores (Adjusted *R²* = .280, *F*(5, 2414) = 189.5, *p* < .001).

### Cohort-specific findings

Due to measurement invariance not holding, within-cohort analyses were run. Cohort-specific analyses revealed both consistent and divergent patterns in the relationship between early life adversity and internalizing and externalizing behaviours. See Table 4 for the standardized estimates (β) for each model.

**Table 4.**
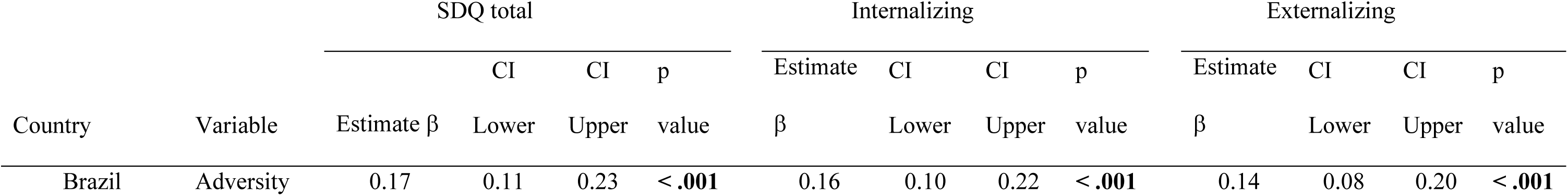
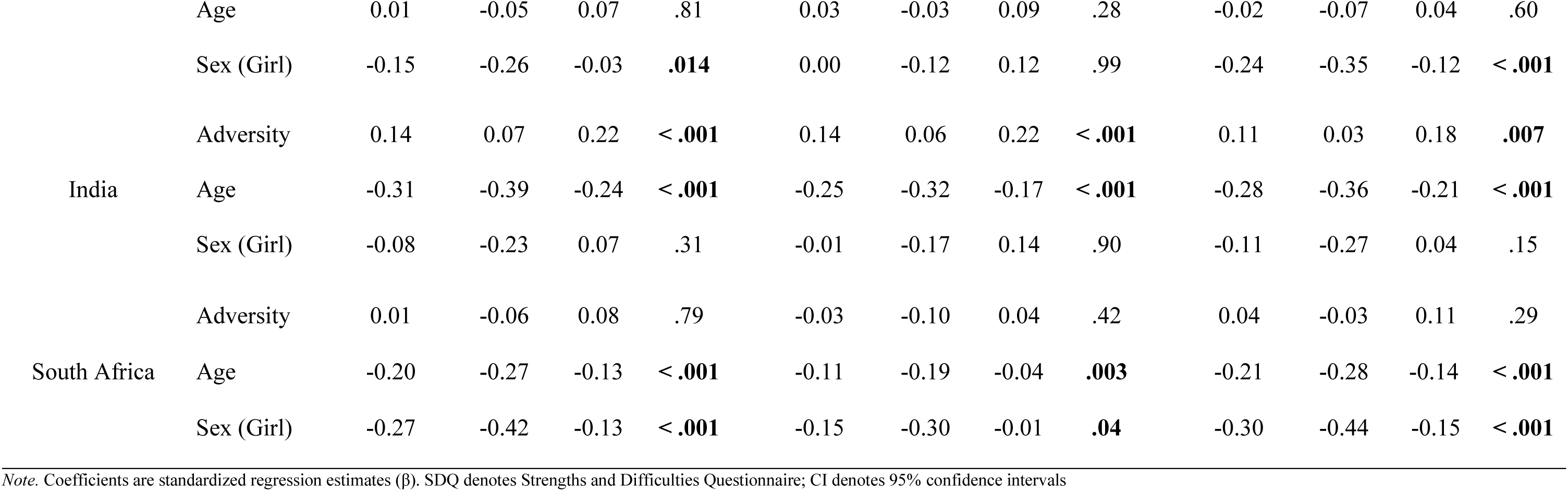
Cohort-specific findings.

| Country | Variable | SDQ total |  |  |  | Internalizing |  |  |  | Externalizing |  |  |  |
| --- | --- | --- | --- | --- | --- | --- | --- | --- | --- | --- | --- | --- | --- |
| | | Estimate $\beta$ | CI | | p value | Estimate $\beta$ | CI | | p value | Estimate $\beta$ | CI | | p value |
|  |  |  | Lower | Upper |  |  | Lower | Upper |  |  | Lower | Upper |  |
| Brazil | Adversity | 0.17 | 0.11 | 0.23 | < .001 | 0.16 | 0.10 | 0.22 | < .001 | 0.14 | 0.08 | 0.20 | < .001 |
|  | Age | 0.01 | -0.05 | 0.07 | .81 | 0.03 | -0.03 | 0.09 | .28 | -0.02 | -0.07 | 0.04 | .60 |
|  | Sex (Girl) | -0.15 | -0.26 | -0.03 | **.014** | 0.00 | -0.12 | 0.12 | .99 | -0.24 | -0.35 | -0.12 | **< .001** |
|  | Adversity | 0.14 | 0.07 | 0.22 | **< .001** | 0.14 | 0.06 | 0.22 | **< .001** | 0.11 | 0.03 | 0.18 | **.007** |
| India | Age | -0.31 | -0.39 | -0.24 | **< .001** | -0.25 | -0.32 | -0.17 | **< .001** | -0.28 | -0.36 | -0.21 | **< .001** |
|  | Sex (Girl) | -0.08 | -0.23 | 0.07 | .31 | -0.01 | -0.17 | 0.14 | .90 | -0.11 | -0.27 | 0.04 | .15 |
|  | Adversity | 0.01 | -0.06 | 0.08 | .79 | -0.03 | -0.10 | 0.04 | .42 | 0.04 | -0.03 | 0.11 | .29 |
| South Africa | Age | -0.20 | -0.27 | -0.13 | **< .001** | -0.11 | -0.19 | -0.04 | **.003** | -0.21 | -0.28 | -0.14 | **< .001** |
|  | Sex (Girl) | -0.27 | -0.42 | -0.13 | **< .001** | -0.15 | -0.30 | -0.01 | **.04** | -0.30 | -0.44 | -0.15 | **< .001** |

*Note.* Coefficients are standardized regression estimates ( $\beta$ ). SDQ denotes Strengths and Difficulties Questionnaire; CI denotes 95% confidence intervals

In BHRC, early life adversity was strongly associated with higher SDQ scores (B = 6.78, 95% CI: 4.49, 9.07, *p* < .001). Girls had lower SDQ scores than boys (B = -1.14, 95% CI: -2.06, -0.23, *p* = .014). In SPRING, adversity also predicted higher SDQ scores (B = 1.54, 95% CI: 0.72, 2.35, *p* < .001). Each additional year of age was associated with lower SDQ scores (B = -3.23, 95% CI: -4.01, -2.45, *p* < .001). In DCHS, adversity was not significantly associated with SDQ scores (B = 0.12, *p* = .78). However, each additional year of age was associated with lower SDQ scores (B = -5.5, 95% CI: -7.50, -3.49, *p* < .001), and girls scored lower than boys (B = -1.23, 95% CI: -1.87, -0.59, *p* < .001).

In BHRC, early life adversity was associated with higher internalizing scores (B = 3.3, 95% CI: 2.10, 4.50, *p* < .001). Neither age nor sex were significant predictors. In SPRING, early life adversity was positively associated with internalizing scores (B = 0.81, 95% CI: 0.36, 1.27, *p* < .001) and each additional year of age predicted lower scores (B = – 1.37, 95% CI: –1.81, –0.94, *p* < .001). In DCHS, early life adversity was not significantly associated with internalizing scores (B = –0.18, *p* = .42). However, both age (B = –1.62, 95% CI: –2.68, –0.56, *p* = .003) and female sex (B = –0.35, 95% CI: –0.70, –0.01, *p* = .041) were associated with lower scores.

In BHRC, early life adversity was associated with higher externalizing scores (B = 3.48, 95% CI: 2.05, 4.91, *p* < .001). Girls had significantly lower scores than boys (B = – 1.15, 95% CI: –1.72, –0.58, *p* < .001). In SPRING, early life adversity was significantly associated with externalizing scores (B = 0.73, 95% CI: 1.19, 1.26, *p* = .007) and each additional year of age predicted lower scores (B = –1.86, 95% CI: –2.36, –1.35, *p* < .001). In DCHS, early life adversity was not significantly associated with externalizing scores (B = 0.3, *p* = .29). However, both age (B = –3.87, 95% CI: –5.19, –2.56, *p* < .001) and female sex (B = –0.88, 95% CI: –1.3, –0.45, *p* < .001) were associated with lower scores.

### Secondary analyses

#### Early life adversity and age interactions

Linear regressions were conducted to examine the association between child age (centered), child sex, cohort, and the interaction of early life adversity and child age (centered), in predicting child behavior in each cohort.

The overall model for SDQ total score was significant (*F*(6, 2413) = 232.2, *p* < .001). There was a significant main effect of early life adversity (B = 2.39, 95% CI: 1.56, 3.23, *p* < .001) and child age (B = -3.01, 95% CI: -4.44, -1.57, *p* < .001). Girls scored lower than boys on SDQ total score (B = -0.95, 95% CI: -1.44, -0.47, *p* < .001). SPRING and DCHS participants had significantly lower SDQ total scores (SPRING: B = -10.81, 95% CI: -11.47, -10.15, *p* < .001; DCHS: B = -6.13, 95% CI: -6.76, -5.5, *p* < .001). There was a significant early life adversity × child age interaction (B = 2.87, 95% CI: 1.4, 4.34, *p* < .001), which can be seen in Figure 2.

**Figure 2.**
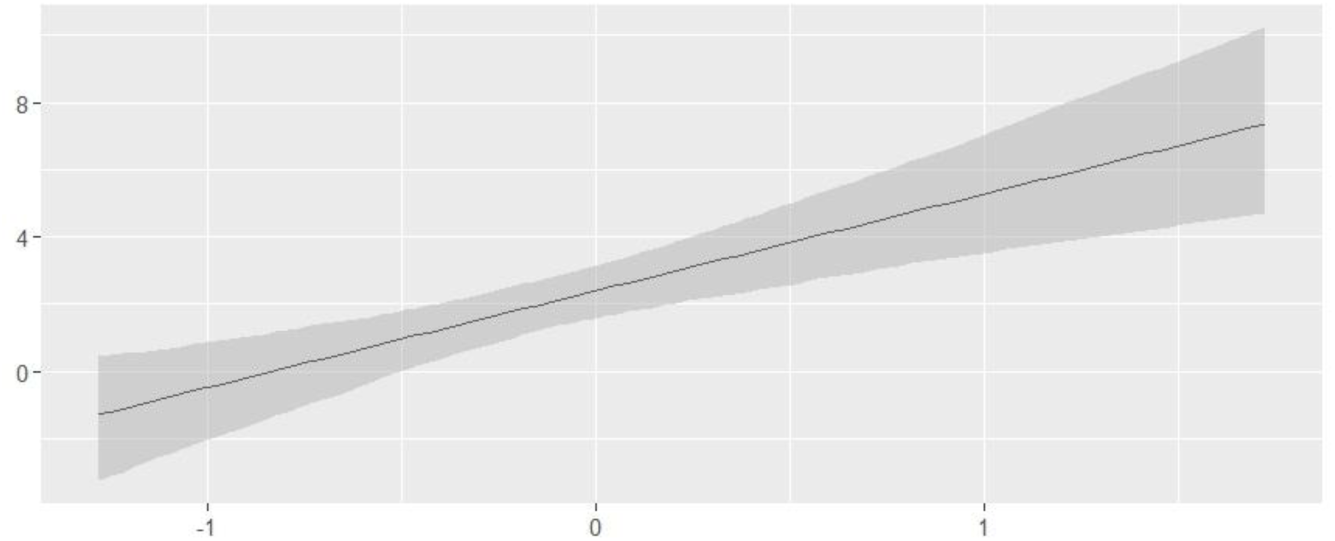
Early life adversity × child age interaction for SDQ total scores.

The overall model for internalizing behavior was significant (*F*(6, 2413) = 208.9, *p* < .001). There was a significant main effect of early life adversity (B = 1.03, 95% CI: 0.59, 1.47, *p* < .001) and child age (B = -1.21, 95% CI: -1.96, -0.45, *p* = .002). SPRING and DCHS participants had significantly lower internalizing behavior scores (SPRING: B = -4.98, 95% CI: -5.33, -4.63, *p* < .001; DCHS: B = -3.83, 95% CI: -4.16, -3.50, *p* < .001). There was a significant early life adversity × child age interaction (B = 1.29, 95% CI: 0.51, 2.06, *p* = .001), which can be seen in Figure 3.

**Figure 3.**
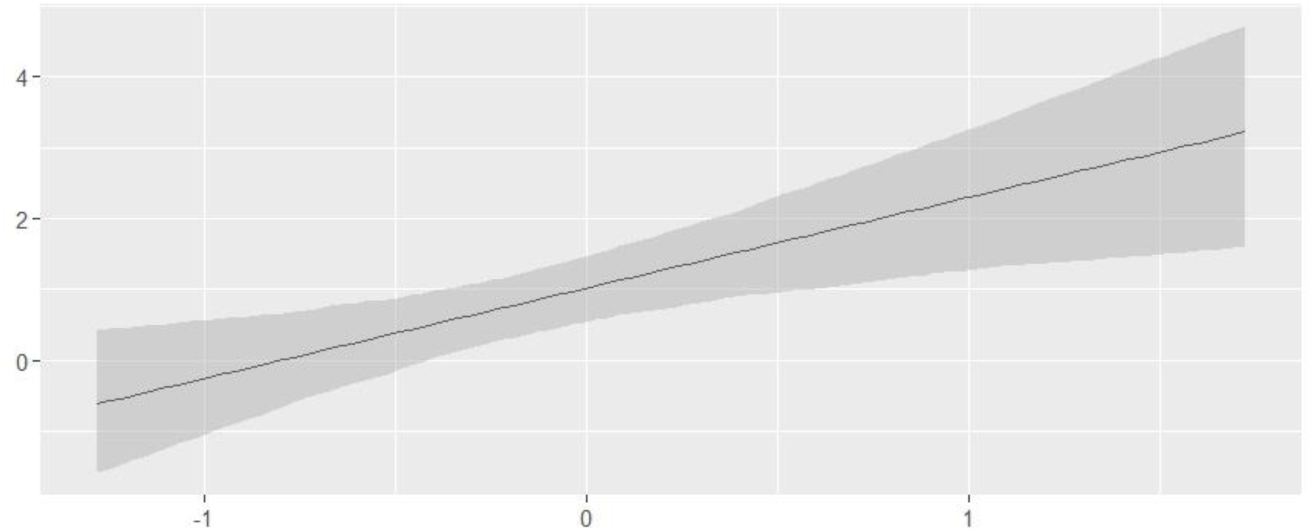
Early life adversity × child age interaction for internalizing scores.

The overall model for externalizing behavior was significant (*F*(6, 2413) = 160.5, *p* < .001). There was a significant main effect of early life adversity (B = 1.36, 95% CI: 0.84, 1.89, *p* < .001) and child age (B = -1.8, 95% CI: -2.7, -0.89, *p* = .006). Girls scored lower than boys on externalizing behaviors (B = -0.85, 95% CI: -1.15, -0.55, *p* < .001). SPRING and DCHS participants had significantly lower externalizing behavior scores (SPRING: B = - 5.83, 95% CI: -6.24, -5.41, *p* < .001; DCHS: B = -2.30, 95% CI: -2.7, -1.91, *p* < .001). There was a significant early life adversity × child age interaction (B = 1.59, 95% CI: 0.66, 2.51, *p* < .001), which can be seen in Figure 4.

**Figure 4.**
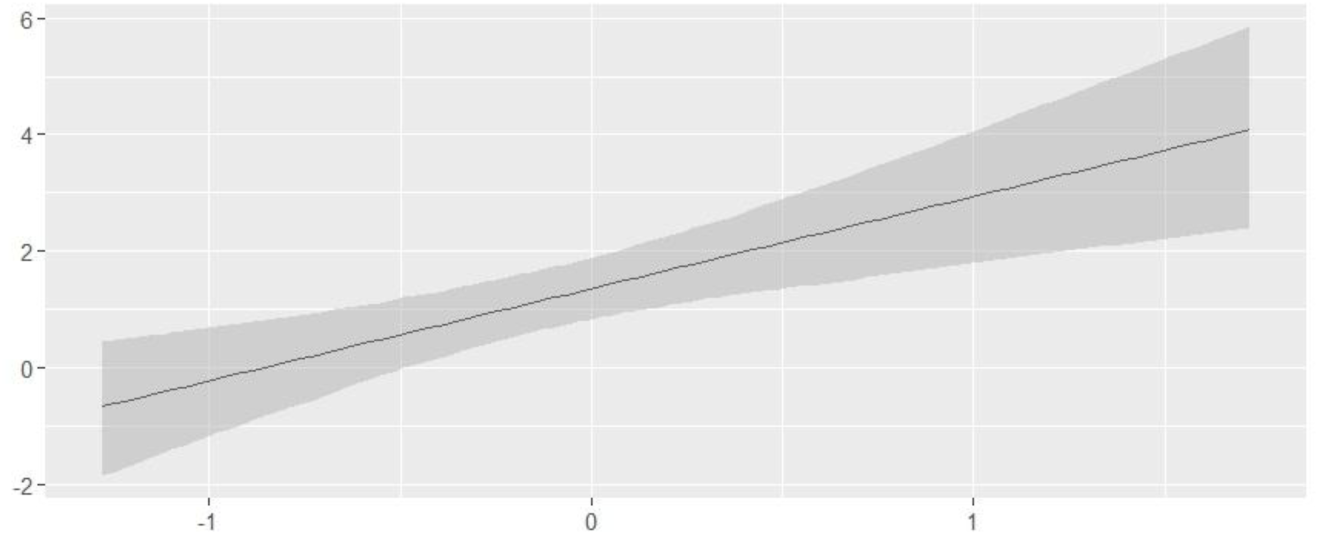
Early life adversity × child age interaction for externalizing scores.

#### Early life adversity and cohort interactions

Linear regressions were conducted to examine the association between age, sex, cohort, and the interaction between early life adversity and cohort, on child behavior.

The overall model for SDQ total scores was significant (*F*(7, 2412) = 203.7, *p* < .001). There was a significant main effect of early life adversity (B = 6.76, 95% CI: 4.97, 8.54, *p* < .001) and child sex, with girls showing lower SDQ total scores than boys (B = - 0.99, 95% CI: -1.47, -0.51, *p* < .001). Compared to BHRC, SPRING had lower scores (B = −5.75, 95% CI: -8.01, -3.49, *p* < .001), while DCHS did not differ significantly (B = −0.2, *p* = .85). There was a significant early life adversity × country interaction, and the effect of early life adversity was significantly reduced in SPRING and DCHS participants, relative to BHRC participants (SPRING: B = -5.29, 95% CI: -7.62, -2.97, *p* < .001; DCHS: B = -6.43, 95% CI: -8.54, -4.31, *p* < .001). Figure 5 highlights this interaction.

**Figure 5.**
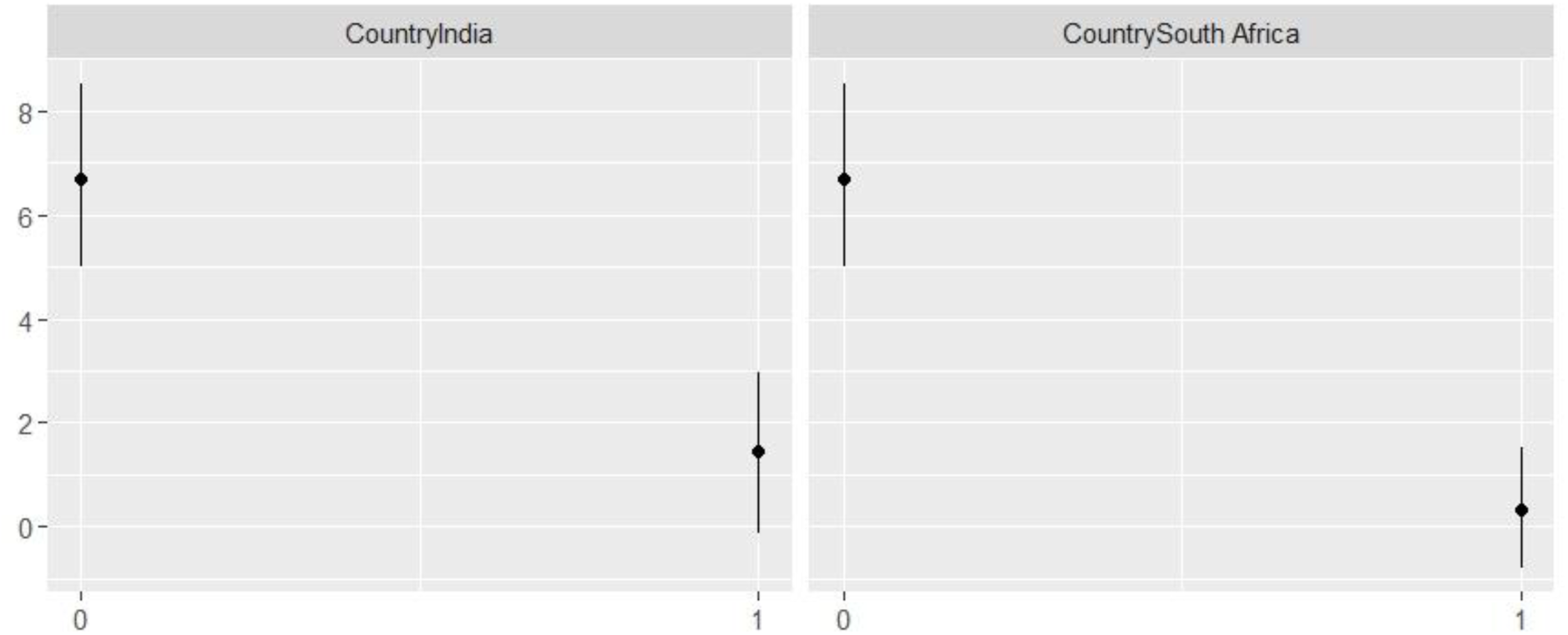
Early life adversity × cohort interaction for SDQ total scores.

The overall model for internalizing behaviors was significant (*F*(7, 2412) = 184.5, *p* < .001). There was a significant main effect of early life adversity (B = 3.31, 95% CI: 2.37, 4.25, *p* < .001). Compared to BHRC, SPRING exhibited lower internalizing scores (B = - 2.53, 95% CI: -3.72, -1.34, *p* < .001), while DCHS did not differ significantly (B = -0.7, *p* = .21). There was a significant early life adversity × country interaction, and the effect of early life adversity was significantly reduced in SPRING and DCHS participants, relative to BHRC participants (SPRING: B = -2.55, 95% CI: -3.77, -1.33, *p* < .001; DCHS: B = -3.41, 95% CI: -4.53, -2.30, *p* < .001). Figure 6 highlights this interaction.

**Figure 6.**
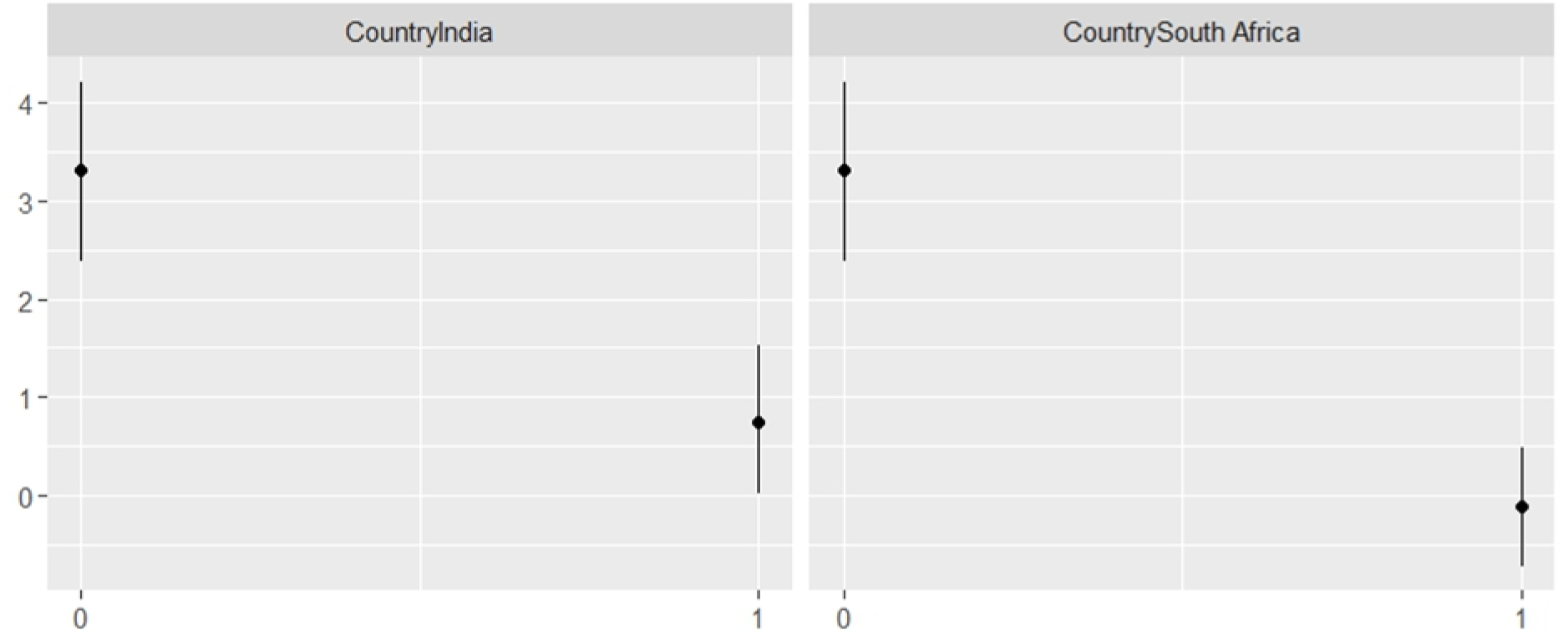
Early life adversity × cohort interaction for internalizing scores.

The overall model for externalizing behaviors was significant (*F*(7, 2412) = 139.4, *p* < .001). There was a significant main effect of early life adversity (B = 3.45, 95% CI: 2.33, 4.58, *p* < .001) and child age (B = -0.31, 95% CI: -0.56, -0.06, *p* = .02). There was a main effect for child sex, with girls showing fewer externalizing behaviors than boys (B = -0.87, 95% CI: -1.17, -0.56, *p* < .001). Compared to BHRC, SPRING exhibited lower externalizing scores (B = -3.22, 95% CI: -4.64, -1.79, *p* < .001), while DCHS did not differ significantly (B = 0.5, *p* = .46). There was a significant early life adversity × country interaction, and the effect of early life adversity was significantly reduced in SPRING and DCHS participants, relative to BHRC participants (SPRING: B = -2.74, 95% CI: -4.21, -1.28, *p* < .001; DCHS: B = -3.01, 95% CI: -4.35, -1.68, *p* < .001). Figure 7 highlights this interaction.

**Figure 7.**
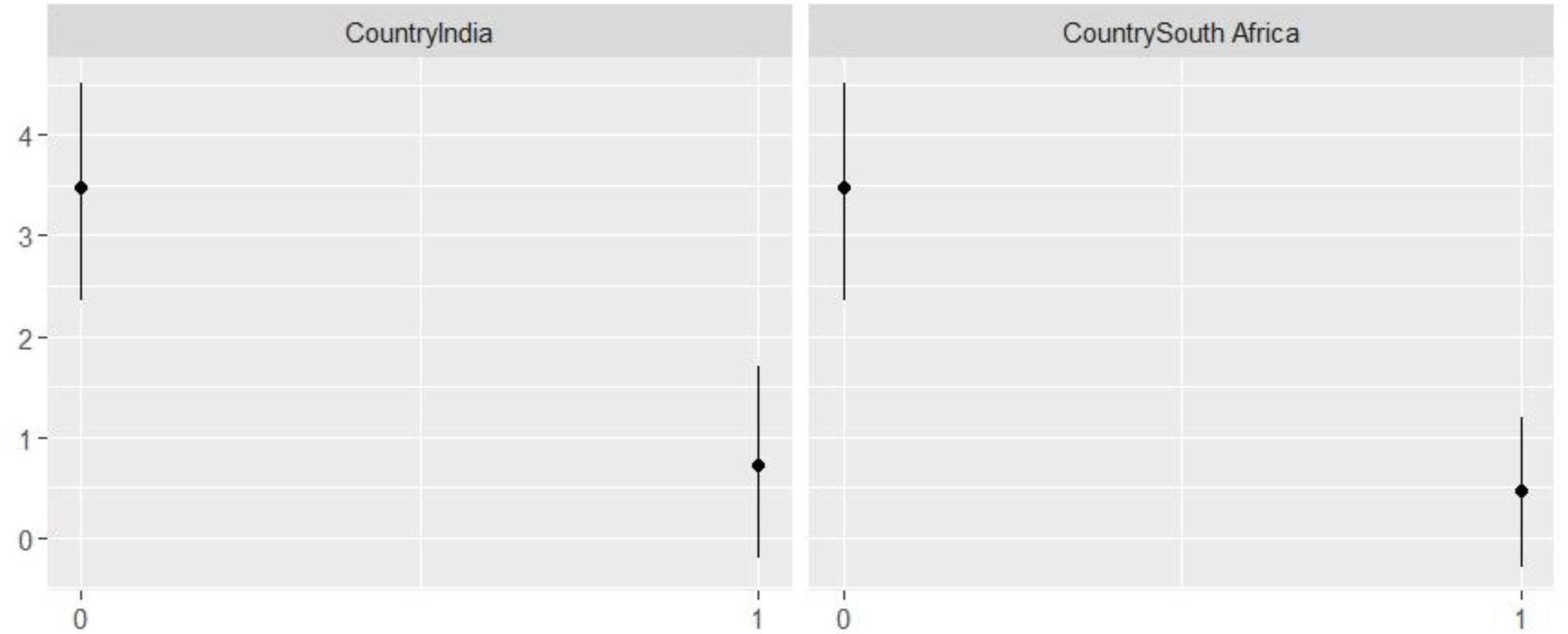
Early life adversity × cohort interaction for externalizing scores.

## Discussion

In pooled and cohort-specific analyses, cumulative ELA in the first two years of life was associated with higher SDQ total, internalizing, and externalizing scores at age 7–10. The strength of these associations varied by cohort (strongest in BHRC, attenuated or non-significant in SPRING and DCHS, respectively), indicating context-dependent effects. Finally, ELA × age interactions suggested larger effects at younger ages in the pooled sample, and sex differences were observed in some cohorts.

Our finding that early life adversity was associated with higher behavioral scores for the SDQ total score, internalizing, and externalizing behaviors is consistent with prior work in high-income countries (Bevilacqua et al., 2021; Haahr-Pedersen et al., 2021). This includes research on translational models (Maccari et al., 2014), clinical cohorts, and community cohorts (Appleyard et al., 2005; Esteves et al., 2017; Vaidya et al., 2024). Large high-income cohorts have reported that multiple childhood adversities were associated with greater internalizing and externalizing behaviors in mid-childhood (Bevilacqua et al., 2021; Haahr-Pedersen et al., 2021). Thus, the association between early life adversity and psychopathology is already evident in mid-childhood.

Cohort-specific findings were observed when the sample was stratified by cohort. Our findings indicated that BHRC participants exhibited a greater association of ELA with behavior outcomes relative to the SPRING cohort, where age effects were observed. The DCHS cohort did not exhibit an association between ELA and behavior outcomes but instead highlighted that child age and sex were predictors of behavioral outcomes. These differences may highlight contextual risk factors (Yoon et al., 2024), or may reflect variation in severity of early life adversity exposure or cultural expression and reporting of child behavior.

We found that the association of early life adversity with behavior is stronger at younger ages. This is consistent with a prior literature that discusses the development of resilience over time (Lee et al., 2025; Miner & Clarke-Stewart, 2008). We found that sex differences differed by country. In the BHRC, girls scored lower than boys on SDQ total score and externalizing behavior, in DCHS girls scored lower than boys on SDQ total score, internalizing, and externalizing behaviors, and in SPRING there were differences in SDQ scores by sex. While sex differences in behavioral outcomes have been observed in the literature, these effects are often small (Carneiro et al., 2016).

Some limitations should be emphasized. First, the adversity variable was heterogeneous between cohorts, therefore, to mitigate heterogeneity in the adversity variable, the adversity variable was dichotomized. Dichotomization may have reduced variability and adversity severity analyses could not be run. Second, this study examined adversity experienced in the first two years of life and adversity experienced after 2 years of age was not included. Third, measurement invariance did not hold across cohorts, indicating that results may not be directly comparable, therefore cohort differences should be interpreted cautiously.

In conclusion, in cohorts from the Global South, while there is an association between early life adversity and psychopathology, there are also important context-dependent differences across cohorts. Early life trauma may be particularly important to identify, and culturally tailored interventions may be needed. Future work should use harmonized measures and longitudinal designs to clarify mechanisms and causal timing.

## Data Availability

All data produced in the present study are available upon reasonable request to the authors

## Acknowledgments

We thank the mothers and their children for participating in the study and the study staff, the clinical and administrative staff of the Western Cape Government Health Department at Paarl Hospital and at the clinics for support of the study. We thank Mauricio Scopel Hoffmann for his guidance and knowledge regarding measurement invariance. His support and code are greatly appreciated. We thank the SPRING and COINCIDE consortia, participating cohort families and the National Health Mission of Haryana for their support.

## Funding

BHRC - The study was funded by Conselho Nacional de Desenvolvimento Científico e Tecnológico (CNPq grant numbers 573974/2008-0 and 465550/2014-2), Fundação de Amparo à Pesquisa do Estado de São Paulo (FAPESP grant numbers: 2008/57896-8, 2013/08531-5, 2014/50917-0, 2020/06172-1, 2021/05332-8, 2021/12901-9), European Research Council (ERC grant numbers: 337673 and 101057390), UK Medical Research Council (MRC grant number: MR/R022763/1), Ministério da Saúde (Decit/SECTICS/MS Grant number: 888379/2019 - Portaria N° 1.949, 04/08/2020) and Banco Industrial do Brasil S/A (CISM grant). Scholarship received financial support from Conselho Nacional de Desenvolvimento Científico e Tecnológico (CAPES). Collaboration between the BHRC and other cohorts has been funded by the National Institutes of Health (NIMH grant number: R01MH120482-01). Involvement of NIMH Intramural investigators has been funded by NIMH-Intramural Research Program Project MH 002782.

SPRING cohort was established through a Wellcome Trust programme grant [093615]; the adversity data was collected through the SPRING-ELS sub-study [107818]; the SDQ data was collected through the COINCIDE study, funded by a Team Science Grant from DBT/Wellcome Trust India Alliance [IA/TSG/20/1/600023].

DCHS - The core study has been funded by the Gates Foundation (OPP1017641, OPP1017579). Additional support for HJZ and DJS was provided by the Medical Research Council of South Africa.

## Notes

### Competing Interest Statement

The authors have declared no competing interest.

### Funding Statement

Funding
BHRC - The study was funded by Conselho Nacional de Desenvolvimento Cientifico e Tecnologico (CNPq grant numbers 573974/2008-0 and 465550/2014-2), Fundacao de Amparo a Pesquisa do Estado de S&atildeo Paulo (FAPESP grant numbers: 2008/57896-8, 2013/08531-5, 2014/50917-0, 2020/06172-1, 2021/05332-8, 2021/12901-9), European Research Council (ERC grant numbers: 337673 and 101057390), UK Medical Research Council (MRC grant number: MR/R022763/1), Ministerio da Saude (Decit/SECTICS/MS Grant number: 888379/2019 - Portaria No. 1.949, 04/08/2020) and Banco Industrial do Brasil S/A (CISM grant). Scholarship received financial support from Conselho Nacional de Desenvolvimento Cientifico e Tecnologico (CAPES). Collaboration between the BHRC and other cohorts has been funded by the National Institutes of Health (NIMH grant number: R01MH120482-01). Involvement of NIMH Intramural investigators has been funded by NIMH-Intramural Research Program Project MH 002782.
SPRING cohort was established through a Wellcome Trust programme grant [093615]; the adversity data was collected through the SPRING-ELS sub-study [107818]; the SDQ data was collected through the COINCIDE study, funded by a Team Science Grant from DBT/Wellcome Trust India Alliance [IA/TSG/20/1/600023].
DCHS - The core study has been funded by the Gates Foundation (OPP1017641, OPP1017579). Additional support for HJZ and DJS was provided by the Medical Research Council of South Africa.

### Author Declarations

Faculty of Health Science's Human Research Ethics committee of University of Cape Town gave ethical approval for this work (HREC 640/2024)

## References

Allen, L., Williams, J., Townsend, N., Mikkelsen, B., Roberts, N., Foster, C., & Wickramasinghe, K. (2016). Poverty and risk factors for non-communicable diseases in developing countries: a systematic review. The lancet, 388, S17.

Appleyard, K., Egeland, B., van Dulmen, M. H., & Alan Sroufe, L. (2005). When more is not better: The role of cumulative risk in child behavior outcomes. Journal of Child Psychology and Psychiatry, 46(3), 235–245.

Berman, I. S., McLaughlin, K. A., Tottenham, N., Godfrey, K., Seeman, T., Loucks, E., Suomi, S., Danese, A., & Sheridan, M. A. (2022). Measuring early life adversity: A dimensional approach. Development and Psychopathology, 34(2), 499–511.

Bevilacqua, L., Kelly, Y., Heilmann, A., Priest, N., & Lacey, R. E. (2021). Adverse childhood experiences and trajectories of internalizing, externalizing, and prosocial behaviors from childhood to adolescence. Child abuse & neglect, 112, 104890.

Bhopal, S., Roy, R., Verma, D., Kumar, D., Avan, B., Khan, B., Gram, L., Sharma, K., Amenga-Etego, S., Panchal, S. N., Soremekun, S., Divan, G., Kirkwood, B. R., & Yousafzai, A. K. (2019). Impact of adversity on early childhood growth & development in rural India: Findings from the early life stress sub-study of the SPRING cluster randomised controlled trial (SPRING-ELS). PloS one, 14(1), e0209122. 10.1371/journal.pone.0209122

Carneiro, A., Dias, P., & Soares, I. (2016). Risk factors for internalizing and externalizing problems in the preschool years: Systematic literature review based on the child behavior checklist 1½–5. Journal of Child and Family Studies, 25, 2941–2953.

Cronholm, P. F., Forke, C. M., Wade, R., Bair-Merritt, M. H., Davis, M., Harkins-Schwarz, M., Pachter, L. M., & Fein, J. A. (2015). Adverse childhood experiences: Expanding the concept of adversity. American journal of preventive medicine, 49(3), 354–361.

Esteves, K., Gray, S. A., Theall, K. P., & Drury, S. S. (2017). Impact of physical abuse on internalizing behavior across generations. Journal of Child and Family Studies, 26(10), 2753–2761.

Goodman, A., Lamping, D. L., & Ploubidis, G. B. (2010). When to use broader internalising and externalising subscales instead of the hypothesised five subscales on the Strengths and Difficulties Questionnaire (SDQ): data from British parents, teachers and children. Journal of Abnormal Child Psychology, 38, 1179–1191.

Goodman, R. (1997). The Strengths and Difficulties Questionnaire: A Research Note Journal of Child Psychology and Psychiatry, 38, 581–586. 10.1111/j.1469-7610.1997.tb01545.x

Goodman, R. (2001). Psychometric Properties of the Strengths and Difficulties Questionnaire. Journal of the American Academy of Child & Adolescent Psychiatry., 40(11), 1337–1345. 10.1097/00004583-200111000-00015

Haahr-Pedersen, I., Hyland, P., Hansen, M., Perera, C., Spitz, P., Bramsen, R. H., & Vallières, F. (2021). Patterns of childhood adversity and their associations with internalizing and externalizing problems among at-risk boys and girls. Child abuse & neglect, 121, 105272.

Hauser, M. D. (2021). How early life adversity transforms the learning brain. *Mind*, Brain, and Education, 15(1), 35–47.

Holden, G. W., Gower, T., & Chmielewski, M. (2020). Methodological considerations in ACEs research. In Adverse childhood experiences (pp. 161–182). Elsevier.

Kirkwood, B. R., Sikander, S., Roy, R., Soremekun, S., Bhopal, S. S., Avan, B., Lingam, R., Gram, L., Amenga-Etego, S., & Khan, B. (2023). Effect of the SPRING home visits intervention on early child development and growth in rural India and Pakistan: parallel cluster randomised controlled trials. Frontiers in nutrition, 10, 1155763.

Lee, J. O., Duan, L., Constantino-Pettit, A., Yoon, Y., Oxford, M. L., Rose, J., & Cederbaum, J. A. (2025). Does the timing matter? The association between childhood adversity and internalizing and externalizing problems from childhood to adolescence and its sex differences. Child abuse & neglect, 163, 107437.

Lu, C., Black, M. M., & Richter, L. M. (2016). Risk of poor development in young children in low-income and middle-income countries: an estimation and analysis at the global, regional, and country level. The lancet., 4(12), e916–e922. 10.1016/S2214-109X(16)30266-2

Lund, C., Breen, A., Flisher, A. J., Kakuma, R., Corrigall, J., Joska, J. A., Swartz, L., & Patel, V. (2010). Poverty and common mental disorders in low and middle income countries: a systematic review. Social science & medicine, 71(3), 517–528.

Ma, X., Biaggi, A., Sacchi, C., Lawrence, A. J., Chen, P.-J., Pollard, R., Matter, M., Mackes, N., Hazelgrove, K., & Morgan, C. (2022). Mediators and moderators in the relationship between maternal childhood adversity and children’s emotional and behavioural development: a systematic review and meta-analysis. Psychological Medicine, 52(10), 1817–1837.

Maccari, S., Krugers, H. J., Morley-Fletcher, S., Szyf, M., & Brunton, P. (2014). The consequences of early-life adversity: neurobiological, behavioural and epigenetic adaptations. Journal of neuroendocrinology, 26(10), 707–723.

McLaughlin, K. A., Sheridan, M. A., Humphreys, K. L., Belsky, J., & Ellis, B. J. (2021). The value of dimensional models of early experience: Thinking clearly about concepts and categories. Perspectives on Psychological Science, 16(6), 1463–1472.

McLaughlin, K. A., Weissman, D., & Bitrán, D. (2019). Childhood adversity and neural development: A systematic review. Annual review of developmental psychology, 1, 277–312.

Miner, J. L., & Clarke-Stewart, K. A. (2008). Trajectories of externalizing behavior from age 2 to age 9: relations with gender, temperament, ethnicity, parenting, and rater. Developmental psychology, 44(3), 771.

Muris, P., Meesters, C., & van den Berg, F. (2003). The Strengths and Difficulties Questionnaire (SDQ) further evidence for its reliability and validity in a community sample of Dutch children and adolescents. European child & adolescent psychiatry, 12, 1–8.

Oh, D. L., Jerman, P., Silvério Marques, S., Koita, K., Purewal Boparai, S. K., Burke Harris, N., & Bucci, M. (2018). Systematic review of pediatric health outcomes associated with childhood adversity. BMC pediatrics., 18(1). 10.1186/s12887-018-1037-7

R Core Team. (2021). R: A Language and Environment for Statistical Computing. *Vienna, Austria*: *R Foundation for Statistical Computing*. In https://www.R-project.org/

Salum, G. A., Gadelha, A., Pan, P. M., Moriyama, T. S., Graeff-Martins, A. S., Tamanaha, A. C., Alvarenga, P., Krieger, F. V., Fleitlich-Bilyk, B., & Jackowski, A. (2015). High risk cohort study for psychiatric disorders in childhood: rationale, design, methods and preliminary results. International journal of methods in psychiatric research, 24(1), 58–73.

Vaidya, N., Marquand, A. F., Nees, F., Siehl, S., & Schumann, G. (2024). The impact of psychosocial adversity on brain and behaviour: an overview of existing knowledge and directions for future research. Molecular psychiatry, 29(10), 3245–3267.

Viola, T. W., Salum, G. A., Kluwe-Schiavon, B., Sanvicente-Vieira, B., Levandowski, M. L., & Grassi-Oliveira, R. (2016). The influence of geographical and economic factors in estimates of childhood abuse and neglect using the Childhood Trauma Questionnaire: A worldwide meta-regression analysis. Child abuse & neglect, 51, 1–11.

Walker, S. P., Wachs, T. D., Grantham-McGregor, S., Black, M. M., Nelson, C. A., Huffman, S. L., Baker-Henningham, H., Chang, S. M., Hamadani, J. D., & Lozoff, B. (2011). Inequality in early childhood: risk and protective factors for early child development. The lancet, 378(9799), 1325–1338.

Yoon, S., Maguire-Jack, K., Ploss, A., Benavidez, J. L., & Chang, Y. (2024). Contextual factors of child behavioral health across developmental stages. Development and Psychopathology, 36(2), 660–673.

Zar, H., Barnett, W., Myer, L., Stein, D., & Nicol, M. (2015). Investigating the early-life determinants of illness in Africa: the Drakenstein Child Health Study. Thorax, 70(6), 592-594.

